# Cortical activity during narrative discourse production in individuals with post-stroke aphasia and controls measured via functional near-infrared spectroscopy

**DOI:** 10.64898/2026.06.05.26354921

**Authors:** Emily J. Braun, Erin A. Carpenter, Yuanyuan Gao, Meryem A. Yücel, David A. Boas, Swathi Kiran

## Abstract

**Introduction:** Aphasia is an acquired language disorder with a significant negative functional impact. Much of the research on aphasia has focused on word-level language comprehension and production. Further evaluation of discourse-level tasks, both at behavioral and neural levels, will allow for an ecologically valid understanding of the functional implications of language impairment in this population.

**Method:** This study evaluated bilateral frontal, temporal, and parietal cortical activity during computer-based narrative production in 14 young neurotypical individuals, 17 individuals with post-stroke aphasia, and 15 age-matched neurotypical participants using functional near-infrared spectroscopy (fNIRS). Oxygenated hemoglobin (HbO) was measured during narrative production following short video clips and compared to HbO during counting aloud. In addition, behavioral measures quantifying in-task performance were correlated with averaged HbO values.

**Results:** Across primary ROIs (bilateral frontal and temporal), there was greater cortical activity for narrative production than counting aloud. Between group differences were seen in right frontal and left temporal ROIs. In the left temporal ROI, there was a condition-related effect for the young neurotypical group only. In the right frontal ROI, there was a condition-related effect for both the young neurotypical group and the individuals with aphasia. Each group showed different patterns in relationships between cortical activity and discourse performance measures.

**Conclusion:** Overall, young participants showing more consistent condition-related effects for narrative discourse production than individuals with aphasia and age-matched controls. This study shows the potential for fNIRS to evaluate cortical activity for ecologically valid language tasks in individuals with post-stroke aphasia.

## 1. Introduction

Aphasia is a language disorder which affects approximately a third of stroke survivors and which has a significant negative impact on communication and quality of life (Flowers et al., 2016). Further understanding of the cognitive and neural underpinnings of impairment in aphasia could potentially assist researchers and clinicians with developing more targeted and effective assessments and intervention.

Behavioral characterization and evaluation of aphasia have often focused on word retrieval given that anomia is a hallmark feature of aphasia. However, communication ultimately involves discourse (i.e., a unit of language longer than a single sentence; Dipper et al., 2021) and individuals with aphasia present with discourse impairments (Stark, 2019). Evaluation of discourse production allows for better understanding of the functional implications of anomia and other features of language impairment in context (Stark, 2019). Furthermore, identification of discourse production impairment may help to identify individuals with more subtle impairment not identified by standardized assessment relying heavily on word-level evaluation.

There are many different types of discourse production, all of which vary in cognitive-linguistic demand. Discourse can involve one speaker (monologue) or two speakers (dialogue). Monologic discourse specifically can be broken into narrative (i.e., story), procedural (i.e., how to do something), and expositional (informational or descriptive; Stark, 2019; Stark et al., 2021). All of these types of monologic discourse involve a variety of complex cognitive skills and require speakers to successfully convey macrostructural (e.g., organization, topic maintenance) and microstructural (i.e., phonological, lexical-semantic, and syntactic) elements of language in order to communicate their message (Dipper et al., 2021). All three of these discourse types are employed by speakers in everyday life. Given that different components of the language system are variably employed across these types of discourse (Stark, 2019), there is value in evaluating multiple discourse types in order to fully understand discourse skills and identify intervention targets.

In parallel to behavioral characterization of aphasia, studies evaluating the neural underpinnings of aphasia, mostly using functional magnetic resonance imaging (fMRI), have also largely focused on single-word tasks (LaCroix et al., 2021; Wilson & Schneck, 2021). Neuroimaging evidence in healthy individuals supports a dual stream model involving frontal, temporal and parietal regions, in which a left-lateralized dorsal stream supports phonological processing and a more bilateral ventral stream supports lexical-semantic processing (Hickok & Poeppel, 2007). Individuals with aphasia tend to show less left-hemisphere activation than controls (Wilson & Schneck, 2021) presumably as a result of the stroke lesion as well as negative impacts of the stroke lesion on the left-hemisphere language network. Furthermore, there is modest evidence that individuals with aphasia may recruit right-hemisphere homologues of damaged regions during language tasks (LaCroix et al., 2021; Wilson & Schneck, 2021).

Discourse processing and production would presumably involve a similar network of fronto-temporoparietal cortical regions and underlying white matter. However, activation patterns during discourse may vary as a result of the unique cognitive-linguistic demands imposed by comprehension and production of connected speech. Furthermore, there is evidence that activation patterns differ for comprehension vs. production of connected speech (Giglio et al., 2024), indicating that comprehension cannot always act as a proxy for production or general language processing.

While the majority of the existing neuroimaging literature has used functional magnetic resonance imaging (fMRI) to evaluate cortical hemodynamics, functional near-infrared spectroscopy (fNIRS) is another non-invasive method that can capture task-based hemodynamic responses. While fNIRS is not as spatially sensitive as fMRI, there are several positive aspects, including 1) relatively less susceptibility to motion artifacts; 2) lack of acoustic noise; 3) ability for participants to be seated in a clinic environment; and 4) relative ease of repeated measures (Quaresima & Ferrari, 2019).

The neurotypical literature using fNIRS to evaluate cortical activity for language production has largely focused on naming tasks; however, limited studies have evaluated discourse. Specifically, greater frontotemporal activity has been found for structured dialogue relative to rote speech production (Suda et al., 2010) and greater prefrontal activity has been found for discourse production relative to discourse comprehension (Cannizzaro et al., 2016). These results suggest greater cognitive-linguistic demand for formulating verbal discourse than tasks that involve limited or no formulation demands.

While the use of fNIRS has increased in recent years, there are few studies specifically examining cortical activity for language tasks in individuals with aphasia. These few studies have highly variable design, task demand, and patterns of results. Most have evaluated confrontation naming (Chang et al., 2022; Gan et al., 2024; Gilmore et al., 2021; Li et al., 2023, 2024, 2026; Sakatani et al., 1998; Xu et al., 2025) or repetition (Hara et al., 2015; Li et al., 2023, 2024), with limited examination of discourse-level production (Braun et al., 2026; Sakatani et al., 1998). Most relevant to the current investigation, Braun and colleagues evaluated cortical activity in individuals with aphasia and control participants during two structured conversational tasks which involved dialogue. This study found, in part, that young control participants tended to show robust, bilateral frontotemporal cortical activity during dialogue that was not seen to the same extent in older control participants and individuals with aphasia. This underscores the importance of considering both age-related and stroke-related effects in evaluating cortical functioning in adults with post-stroke aphasia. At the neural level, individuals with aphasia may be experiencing stroke-related maladaptive or compensatory mechanisms on top of age-related effects on cortical function such as dedifferentiation (Koen & Rugg, 2019).

Further investigation of cortical activity vis fNIRS during a variety of language tasks in people with aphasia and controls, including different types of monologic discourse production (i.e., narrative, procedural, and expositional) relevant to everyday communication would provide additional information about task demand at the neural level. This would allow us to characterize underlying neural mechanisms supporting (or interfering with) successful discourse production in post-stroke aphasia.

The purpose of the current study is to evaluate cortical activity for monologic narrative discourse production in individuals with aphasia as well as neurotypical control participants using fNIRS and relate this activity to behavioral performance.

Research Questions:

1. Across three group of participants (young neurotypical, age-matched neurotypical, individuals with aphasia), are there differences in cortical activity for narrative production (experimental task) vs. counting aloud (control task) in bilateral fronto-temporoparietal ROIs as measured via fNIRS?

It is hypothesized that in bilateral frontotemporal ROIs that there will be a significant condition by group interaction such that a) there is greater cortical activity for narrative vs. counting across groups reflective of greater cognitive-linguistic demand for narrative production than counting; and b) there is greater cortical activity for young neurotypical than age-matched neurotypical and for age-matched neurotypical than people with aphasia overall and in the narrative condition specifically reflective of reduced cortical activity during cognitive-linguistic tasks for older individuals as a result of aging and for those with neural damage following stroke. These hypotheses are supported by previous studies with neurotypical participants showing greater cortical activity for discourse production than speech and language tasks with less formulation demands (Cannizzaro et al., 2016; Suda et al., 2010) and by prior studies showing lower left-hemisphere cortical activity for language tasks in people with aphasia compared to controls (Schneck et al., 2026; Wilson & Schneck, 2021).

Bilateral parietal regions are exploratory ROIs given their known involvement in language processing (Hickok & Poeppel, 2007) but lack of robust evidence for consistent activation trends in the post-stroke aphasia literature (Kiran et al., 2019). It is hypothesized that bilateral precentral gyri will not show by-condition differences given hypothesized similar speech production demands for narrative and counting.

2. In three groups (young neurotypical, age-matched neurotypical, individuals with aphasia), are there relationships between cortical activity during narrative production and in-task behavioral performance in bilateral fronto-temporoparietal ROIs?

It is hypothesized that, for each group, there will be a significant positive relationship between macrostructural and microstructural linguistic measures of online behavioral performance in narrative production and cortical activity in bilateral frontotemporal ROIs and precentral ROIs reflective of a benefit on behavioral performance of greater task-based cortical engagement. This hypothesis is supported by prior literature showing a positive relationship between cortical activity and language task performance in people with aphasia (Schneck et al., 2026; Wilson & Schneck, 2021). As above, bilateral parietal regions are exploratory ROIs.

## 2. Materials and Methods

### 2.1 Participants

The study was approved by the Institutional Review Board at Boston University and all participants provided written informed consent prior to study participation. Eligibility was determined by participant history questionnaires (all participants), review of medical records requested from participants (individuals with aphasia), and behavioral assessments (individuals with aphasia and age-matched neurotypical adults). Participants were 14 young neurotypical adults (mean age = 23.5 years, SD = 4.1), 17 individuals with chronic post-stroke aphasia (mean age = 61.6, SD = 9.2), and 15 age-matched neurotypical adults (mean age = 59.5 years, SD = 9.6). Young neurotypical individuals (6 female; 8 male) were recruited from a university setting (See Table 1 for individual demographics for the young neurotypical group). Three young participants reported a history of depression.

**Table 1:** Young Neurotypical Participant Demographics.

| ID | Age | Sex | Handedness | Race | Ethnicity | Native Language | Other Languages | Years Education |
|---|---|---|---|---|---|---|---|---|
| Y1 |  | male | R | white | NH | English | - | 14 |
| Y2 |  | male | R | white | Hispanic | English | Spanish | 14 |
| Y3 |  | female | R | white | NH | English | Russian | 20 |
| Y4 |  | male | R | white | NH | English | French* | 19 |
| Y5 |  | female | R | Asian | NH | English | Korean | 13 |
| Y6 |  | female | R | white | NH | English | Italian* | 19 |
| Y7 |  | female | R | Asian | NH | English | Korean | 15 |
| Y8 |  | female | R | white | NH | English | - | 18 |
| Y9 |  | male | R | Asian | Hispanic | English | Spanish | 13 |
| Y10 |  | female | R | Black | NH | Igbo | English | 20 |
| Y12 |  | male | R | white | Hispanic | English | Spanish | 16 |
| Y13 |  | male | Ambidextrous | Black | NH | English | French, Patois | 15 |
| Y14 |  | male | R | Asian | NH | English | Urdu | 18 |
| Y15 |  | male | R | white | Hispanic | English | Spanish, Danish, Japanese | 15 |
| Mean | 23.5 |  |  |  |  |  | *basic | 16.4 |
| SD | 4.2 |  |  |  |  |  |  | 2.6 |

NH: non-Hispanic

Age-matched neurotypical participants (9 female; 6 male) were recruited via contacting individuals from an existing database in the Center for Brain Recovery at Boston University and through self-referral to the Center for Brain Recovery (See Table 2 for individual demographics for the age-matched group). One age-matched participant reported a history of dyscalculia as a child. All age-matched neurotypical participants scored within normal limits on the Mini Metal State Examination, 2^nd^ Edition Expanded Version (MMSE-2; Folstein & Folstein, 2010) as a cognitive screening (average t-score = 56, standard deviation = 6, range = [46, 63]).

**Table 2:**
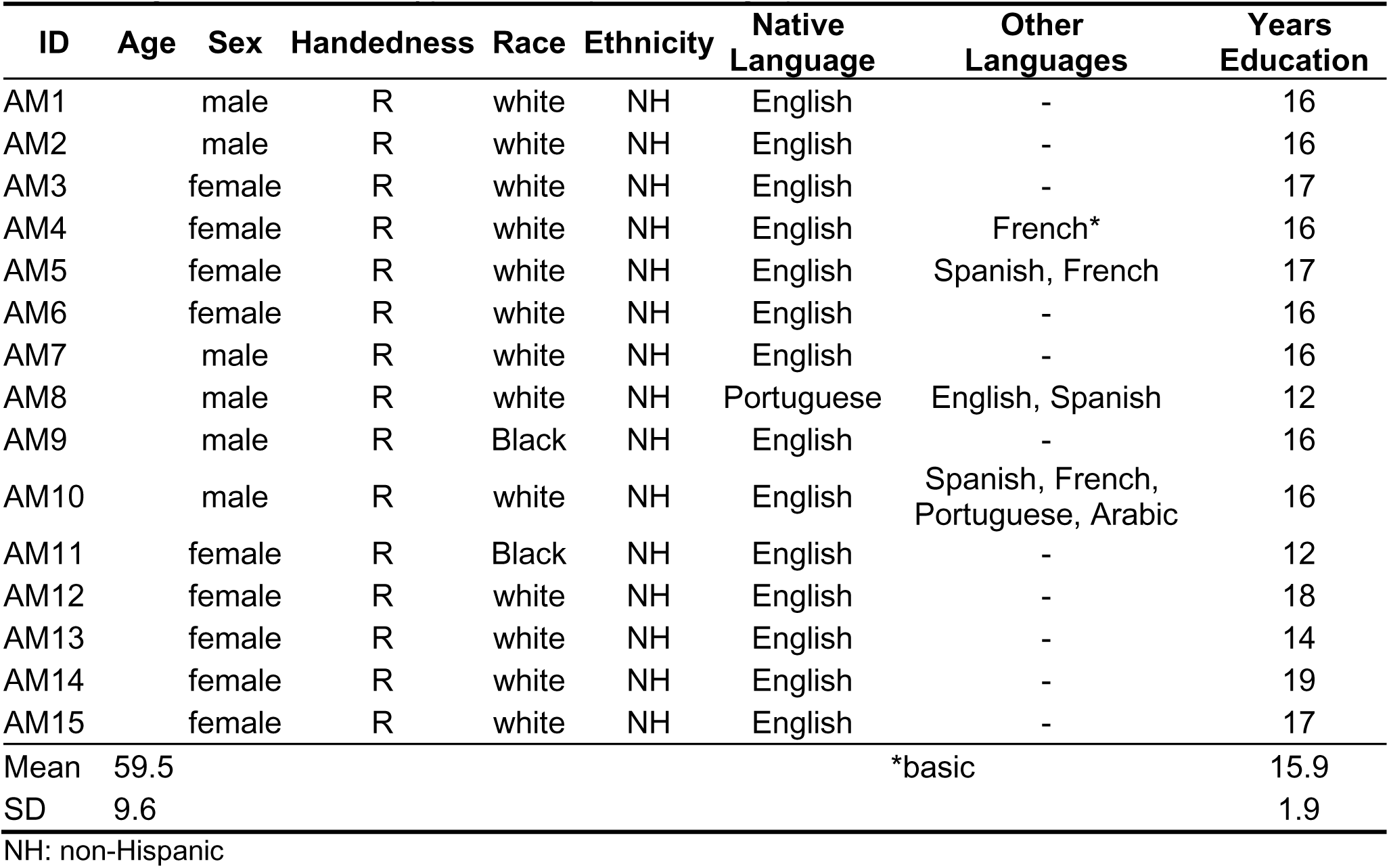
Age-Matched Neurotypical Participant Demographics.

| ID | Age | Sex | Handedness | Race | Ethnicity | Native Language | Other Languages | Years Education |
|---|---|---|---|---|---|---|---|---|
| AM1 |  | male | R | white | NH | English | - | 16 |
| AM2 |  | male | R | white | NH | English | - | 16 |
| AM3 |  | female | R | white | NH | English | - | 17 |
| AM4 |  | female | R | white | NH | English | French* | 16 |
| AM5 |  | female | R | white | NH | English | Spanish, French | 17 |
| AM6 |  | female | R | white | NH | English | - | 16 |
| AM7 |  | male | R | white | NH | English | - | 16 |
| AM8 |  | male | R | white | NH | Portuguese | English, Spanish | 12 |
| AM9 |  | male | R | Black | NH | English | - | 16 |
| AM10 |  | male | R | white | NH | English | Spanish, French, Portuguese, Arabic | 16 |
| AM11 |  | female | R | Black | NH | English | - | 12 |
| AM12 |  | female | R | white | NH | English | - | 18 |
| AM13 |  | female | R | white | NH | English | - | 14 |
| AM14 |  | female | R | white | NH | English | - | 19 |
| AM15 |  | female | R | white | NH | English | - | 17 |
| Mean | 59.5 |  |  |  |  |  | *basic | 15.9 |
| SD | 9.6 |  |  |  |  |  |  | 1.9 |

NH: non-Hispanic

Individuals with aphasia (4 female; 13 male) were recruited from an existing database in the Center for Brain Recovery at Boston University as well as through contacting local clinicians, aphasia groups, and community groups (see Table 3 for individual demographics for the individuals with aphasia). All individuals with aphasia met the following inclusion criteria: 1) chronic aphasia due to a left-hemisphere stroke, 2) mild-moderate aphasia severity (i.e., Western Aphasia Battery – Revised (WAB-R) Aphasia Quotient (AQ) ≥ 50) (Kertesz, 2007) given the task demand, and 3) no significant neurological or psychiatric history prior to stroke. A pure tone audiometric screening was performed at 30 dB HL at 500, 1000, 2000, and 4000 Hz for the individuals with aphasia and the age-matched neurotypical individuals given the possibility of age-related hearing loss. For the hearing screening, 12/15 age-matched participants and 11/17 individuals with aphasia passed in at least one ear. For individuals who failed the hearing screening, increased volume during all study tasks was offered and the experimenter verified with the participants whether they perceived the volume was adequate throughout administration.

**Table 3:** Participant Demographics for People with Aphasia.

| ID | Age | Sex | Handedness | Race | Other Languages | Years Education | Months Post-Onset | Stroke Location (per medical record) |
|---|---|---|---|---|---|---|---|---|
| A1 |  | male | L | white | - | 17 | 70 | Left MCA (fronto-temporoparietal) |
| A2 |  | male | R | white | Spanish | 19 | 21 | Left MCA (parietal) |
| A3 |  | male | R | Black | - | 12 | 152 | Left MCA infarct (fronto-temporal, subcortical) |
| A4 |  | male | R | white | - | 12 | 162 | Left MCA |
| A5 |  | female | R | white | - | 16 | 26 | Left hemisphere (temporal, subcortical) |
| A6 |  | male | R | white | - | 20 | 84 | Left MCA |
| A7 |  | male | R | Black | - | 12 | 44 | Left MCA (temporal) |
| A8 |  | male | R | white | - | 16 | 57 | Left MCA |
| A9 |  | female | R | white | Spanish | 17 | 154 | Left hemisphere (fronto-parietal) |
| A10 |  | male | L | white | - | 12 | 299 | Left hemisphere |
| A11 |  | female | R | Black | - | 16 | 73 | Left hemisphere* |
| A12 |  | male | R | white | - | 16 | 16 | Left hemisphere (thalamus, basal ganglia) |
| A13 |  | male | R | white | - | 16 | 244 | Left hemisphere (temporoparietal) |
| A14 |  | female | R | white | Spanish | 18 | 8 | Left hemisphere |
| A15 |  | male | R | white | - | 18 | 127 | left MCA |
| A16 |  | male | R | white | - | 12 | 55 | Left hemisphere (basal ganglia) |
| A17 |  | male | R | white | - | 16 | 53 | left MCA |
| Mean | 61.6 |  |  |  |  | 15.6 | 96.8 | *history of multiple strokes |
| SD | 9.2 |  |  |  |  | 2.6 | 82.4 |  |

Ethnicity – all participants were non-Hispanic; Native language – native language English for all participants

### 2.2 Behavioral assessment for individuals with aphasia

Behavioral assessment was completed with individuals with aphasia to determine study eligibility and to characterize their impairment and its functional impact. Assessment included the Western Aphasia Battery Revised Aphasia Quotient (WAB-R AQ; Kertesz, 2007) to measure aphasia severity; Cognitive Linguistic Quick Test – Plus (CLQT+; Helm-Estabrooks, 2017) to measure non-linguistic cognitive skills; Boston Naming Test, 2^nd^ Edition (BNT-2; Kaplan et al., 2001) to measure confrontation naming; Apraxia of Speech Rating Scale v3.5 (ASRS v3.5; Duffy et al., 2023) to evaluate presence and severity of motor speech impairment; the Stroke and Aphasia Quality of Life Scale (SAQOL-39; Hilari et al., 2003) to measures quality of life across physical, communication, and social domains; and the Communicative Effectiveness Inventory (CETI; Lomas et al., 1989), a measure of communication completed by a care partner.

### 2.3 Experimental task design

The task was comprised of two runs of 13 minutes and 16 seconds each. Each run contained four 68-second silent narrative video clips each followed by a 15-second response period and four 68-second non-narrative clips each followed by a 15-second response period (four experimental and four control episodes per run). During the 15-second response period, participants either summarized a narrative video clip (experimental condition) or counted aloud from 1 to 15 after watching a non-narrative video clip (control condition). During the response period, participants saw a progress bar on the screen to allow them to gauge how much time remained. A 15-second baseline with a fixation cross was presented at the beginning and end of each run and between conditions in the run (four blocks of one condition were presented consecutively, followed by four blocks of the other condition, to minimize cognitive switching demands for the individuals with aphasia). After each answer block within a condition, an inter-stimulus interval varying among 5, 10 and 15 seconds was presented (See Figure 1 for task design).

**Figure 1:**
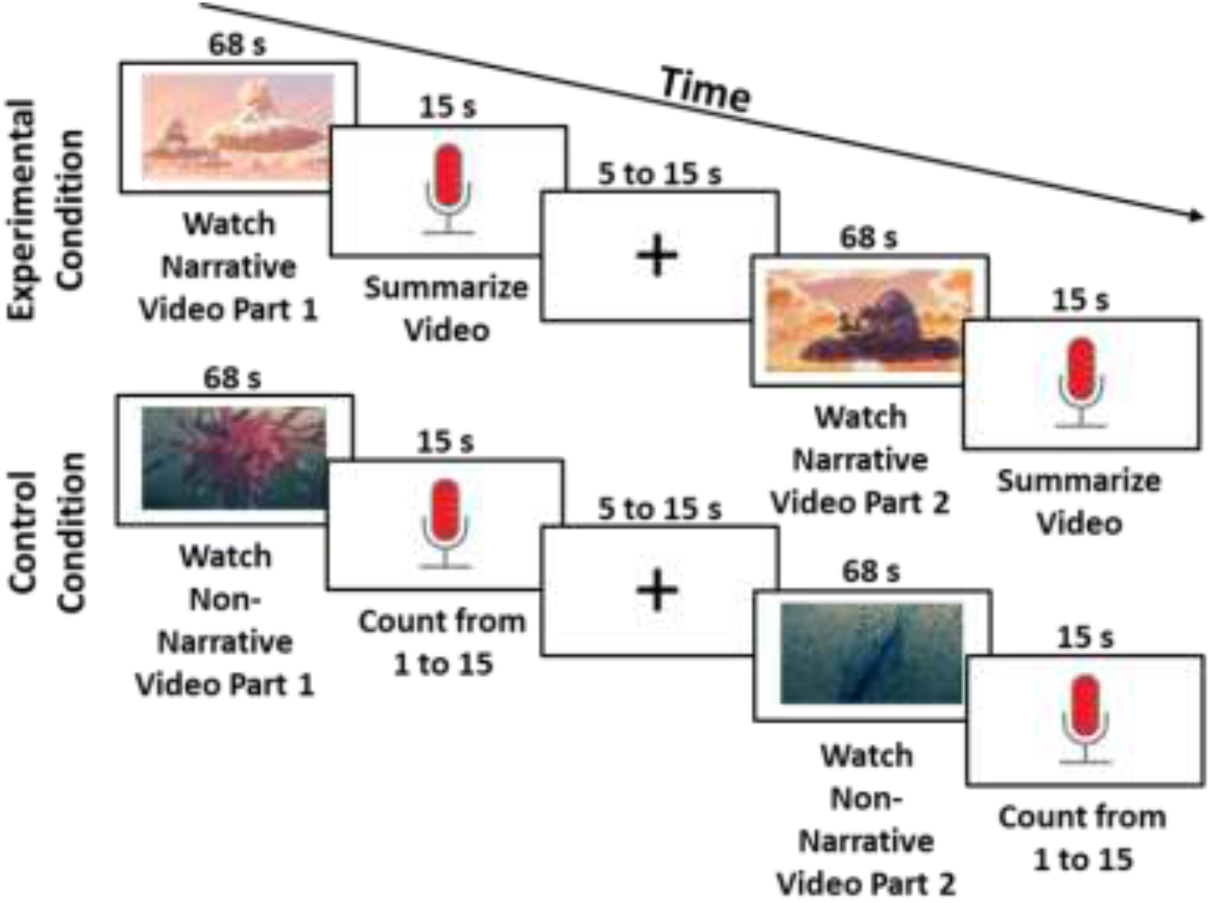
Task Design. Within each of two runs, participants saw four experimental condition blocks and four control condition blocks. In the experimental condition blocks, participants watched narrative video clips in 68-second increments and then were asked to summarize the narrative in a 15-second time period. In the control condition blocks, participants watched a 68-second non-narrative video clip and were asked to count aloud from 1 to 15 in a 15-second time period.

Following the two runs of the task, 10 yes/no questions per silent narrative video set (10 questions for each run) pertaining to the plot were presented via written sentences and corresponding audio recordings. Participants were asked to answer via button press offline (after taking off the fNIRS cap) for each run (20 questions total) to verify attention to the task and comprehension of the story.

To promote comfort with task demands, participants were first provided with verbal instructions prior to completing a practice task. Individuals with aphasia and age-matched participants were additionally provided with a pictorial version of the instructions. Then, a practice task was administered prior to the experiment. Participants were then given an opportunity to ask questions about task completion. The practice task could be completed as many times as necessary until participants were comfortable with the task.

The experiment was first completed with the young neurotypical group and then modified to accommodate individuals with aphasia. Additional visual representation of the experimental conditions was included in the instructions to promote comprehension of task demand. Increased pause time (controlled by the experimenter) was included between conditions in a run to minimize cognitive switching demands. One individual with more severe production impairment was provided with choral cues (i.e., speaking aloud in unison with clinician) during the counting condition as needed to promote greater accuracy.

During the experiment, auditory information was presented using a Jabra speaker and verbal responses were recorded using a Jabra microphone.

### 2.4 Experimental task development

For the stimuli videos, audio was first removed from the Pixar shorts *Partly Cloudy* and *Presto* (Sohn, 2009; Sweetland, 2008). The beginning and end of the videos were cut and then the central portion of the videos was split into four 68-second clips. The clips were presented sequentially so that they followed a logical story sequence. Non-narrative videos were drawn from Inscapes, originally designed for use in fMRI (Vanderwal et al., 2015). Follow-up yes/no comprehension questions were generated by the author and tested with one speaker of American English. Following feedback from this speaker, questions were curated and modified for clarity. Questions were audio-recorded as separate .wav files. Audio editing was done using Apple iMovie software.

The behavioral task was programmed by two research assistants using PsychoPy3 2021.1.2, experimental presentation software that allows for time-locking of experimental events to fNIRS data via LSL triggers (*PsychoPy3*, 2021). This process included creating PsychoPy experiments for two runs of the experimental task and one run of the practice task with use of the edited Pixar short video clips and mp3 audio files (yes/no questions). The programmed experiment included two counterbalance conditions for each of the two runs of the task (four experimental video clips plus responses followed by four control video clips plus responses or vice versa). The behavioral task was piloted with three fluent English speakers. Slight adjustments to timing and computer screen lighting were made based on pilot participant feedback.

### 2.5 Evaluation of task performance

All audio recordings were broadly transcribed in English orthography by the first author and trained research assistants. For the young neurotypical individuals, the first author reviewed 100% of summary condition transcriptions and made corrections as needed. For the individuals with aphasia, the first author reviewed 50-100% of summary condition transcriptions for 9/17 participants at the beginning of and mid-way through the transcription process and provided feedback to the scorer to promote improved consistency and accuracy. The scorer revised transcriptions based on this discussion.

During transcription, the individual transcribing made a manual marking rounded to the nearest second at the beginning and end of the participant response in each block. The talk time annotations at the end of talking in each block were then averaged within each participant to create two values per participant for this experiment (one value for each condition). These values were used as regressors in statistical models to account for varying talk times across conditions and participants.

Transcribed language samples were analyzed with discourse measures at the macrostructural level to evaluate broad planning and organization of discourse as well as at the microstructural level to evaluate lexical-semantic and syntactic aspects of linguistic content. At the language macrostructural level, global coherence, a measure indicating how well the response maintains the overall topic, was evaluated (Harris Wright & Capilouto, 2012).

Transcribed responses were separated into c-units (syntactic unit consisting of an independent clause with all dependent clauses), each given a global coherence rating on an ordinal scale (from 1 (low coherence) to 4 (high coherence)) indexing adherence to overall topic, and then averaged within and across trials to create one averaged global coherence rating for each participant ranging from 1 to 4. The method follows that outlined by Wright and Capilouto (Harris Wright & Capilouto, 2012). One modification was made to the procedure such that utterances that were not a complete independent clause could be considered a separate c-unit if they represented a complete idea.

The coherence ratings were made by the first author and trained research assistants. For the young neurotypical group, a second rater independently completed c-unit separations and global coherence ratings for 60% of the data (9/15 participants). Reliability for averaged coherence ratings was 100%. For the age-matched group, a second rater made c-unit separations and coherence ratings for 27% of the data (4/15 participants). Reliability between the two scorers for averaged coherence ratings was 88%. For the individuals with aphasia, the first author reviewed all c-unit separations and coherence ratings for 35% of the data (6/17 participants) mid-way through completion and then discussed and resolved discrepancies with the rater.

At the language microstructure level, mean length of utterances (MLU) in morphemes, type token ratio (a measure of lexical diversity), and ratio of nouns to verbs (a measure of syntactic complexity), were analyzed. To accomplish this, transcriptions were converted to Computerized Language Analysis (CLAN) format (MacWhinney, 2000) and the mor command was used to generate the linguistic metrics.

### 2.6 Structural MRI

When available for individuals with aphasia, pre-existing T1-weighted structural MRI scans were used, either from previous research participation (N = 11) or from clinical MRI scans (N = 5). For the 11 participants who had completed a previous study in the laboratory, T1-weighted structural MRI scans and corresponding lesion maps drawn using ITK-Snap were available (*ITK-Snap*, 2018). Separately for these 11 participants, head measurements were input into AtlasViewer (Aasted et al., 2015) and estimated individual MNI coordinates for the center of each channel were generated. Following this, channel center coordinates falling within lesioned tissue were identified using MRIcron. Subsequently, channels identified using MRIcron were manually excluded at the participant level in Homer3 prior to running the data processing stream to minimize any potential contribution of spurious signal from the area of lesion (Gilmore et al., 2021). For four participants with clinical MRIs, the scans were reviewed and general lesion location was confirmed. Subsequently, all channels in the lobe(s) of the lesion were manually excluded prior to running the processing stream (see fNIRS data processing). For the remaining participant with a clinical MRI, all left-hemisphere channels were manually excluded given cranial implants that would likely impact signal detection. For the participant without an available research or clinical scan, although clinical records indicated a left-hemisphere stroke, exact lesion location in relation to fNIRS channels was unknown. For this participant, all left-hemisphere channels were excluded.

### 2.7 fNIRS probe design

The fNIRS probe was designed in AtlasViewer (Aasted et al., 2015) to cover bilateral frontal (IFG and MFG), precentral, temporal (STG and MTG), and parietal (AG and SMG) ROIs. This included 36 long channels and eight short separation channels (See Figure 2).

**Figure 2:**
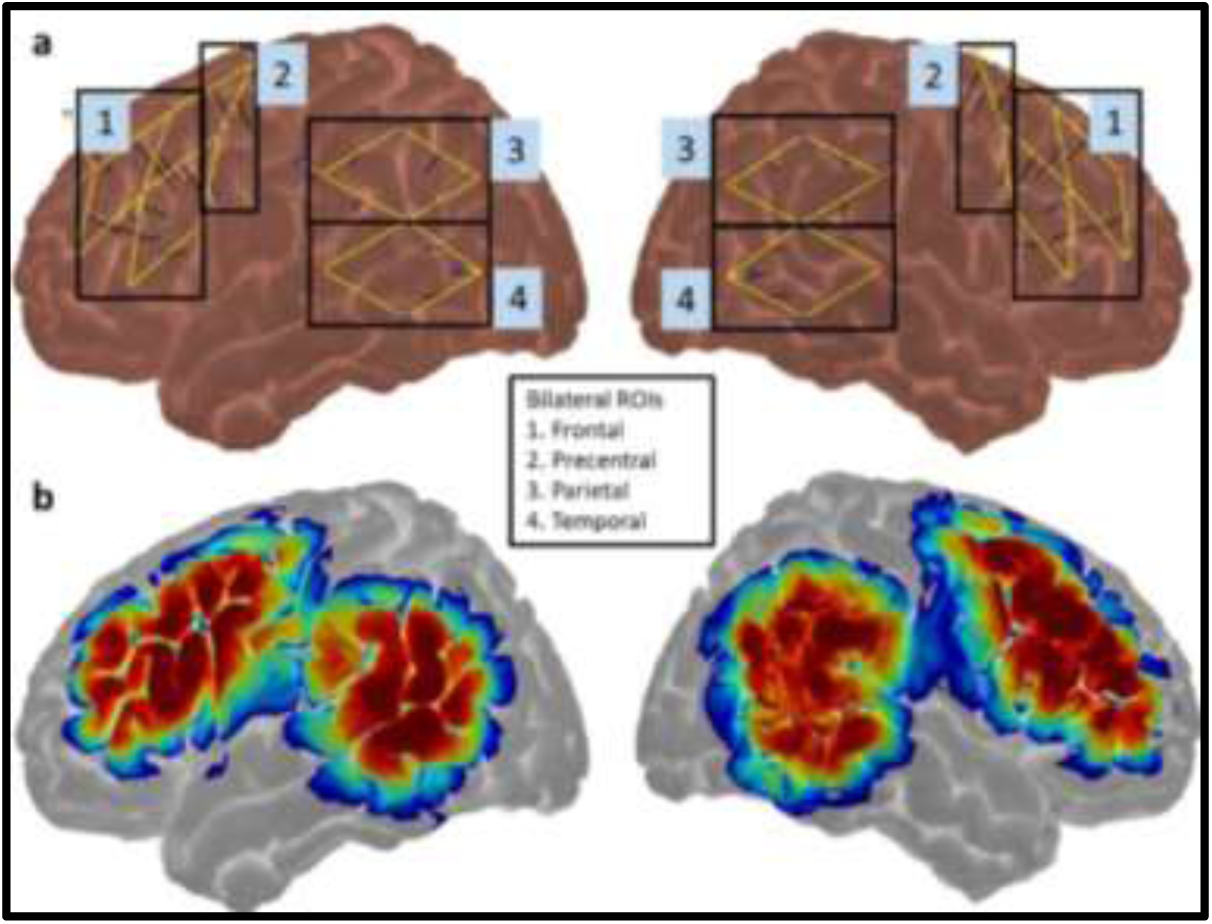
fNIRS Probe Design. Panel (a) indicates bilateral channel location and ROI membership (bilateral frontal, precentral, parietal, and temporal) for each channel. Panel (b) indicates the bilateral sensitivity profile for the probe generated in AtlasViewer (Aasted et al., 2015) with dark red indicating the highest levels of sensitivity.

### 2.8 fNIRS data collection

FNIRS data was collected using a NIRx NIRSport2 (*NIRx NIRSport2*). Data collection took place in a relatively quiet, well-lit room with participants seated at a computer screen. Prior to cap placement, head measurements were taken to ensure proper cap size selection and for use in later analyses. Head measurements included head circumference, distance from nasion to inion, and distance from left preauricular to right preauricular points. Following this, cap placement and adjustment was made to promote the best possible signal quality with adequate scalp to optode coupling. After initial calibration, an opaque shower cap was placed on the participant’s head to reduce effects of ambient light on the detectors. Then, an additional check for signal quality was completed with adjustments as needed. All participants completed a five-minute resting trial prior to the experimental task. In-home assessment was completed with 3/17 participants with aphasia who reported difficulty traveling to the laboratory.

### 2.9 fNIRS data processing

Prior to running the fNIRS processing stream, blocks with no response or responses in error (i.e., participant counted aloud instead of summarizing) were omitted. Total omitted blocks due to no response or counting during the summary condition amounted to 4% of blocks in the individuals with aphasia. The processing stream included 1) pruning of channels with SNR < 5 dB; 2) conversion of raw light intensity data to optical density; 3) motion artifact correction using a spline interpolation method with Savitsky-Golay filtering (frame size = 10 seconds; *p* = .99; Jahani et al., 2018); 4) lowpass filtering to remove high-frequency noise (filter frequency = of 0.5 Hz); 5) conversion of optical density to concentration units using the modified Beer-Lambert Law without path-length correction; and 6) estimation of the hemodynamic response function using a generalized linear model at the run level for changes in HbO and HbR across the block time course with short separation regression (using the short separation channel with the greatest correlation to each channel) and signal drift correction (trange = -2 to 20 seconds; Huppert et al., 2009). See Supplemental Material Figure 1 for processing stream parameters. After running the processing stream, channels that were highly aberrant at the run level (i.e., substantially greater periodicity or amplitude in post-processed data than other channels in that run) were manually excluded and the processing stream was re-run. This included one channel of data for one age-matched participant across both runs due to known equipment malfunction (3% of long channels for this participant).

### 2.10 Statistical analysis

For each participant, channel-level data were averaged across channels into eight broad ROIs: bilateral frontal (IFG and MFG), bilateral PCG, bilateral temporal (MTG and STG) and bilateral parietal (AG and SMG). Statistical analysis and data visualization was completed in R v4.3.2 (R Core Team, 2021) and RStudio (RStudio Team, 2020) with the use of the following packages: tidyverse, dplyr, ggplot2, lme4, and emmeans (Bates et al., 2015; Lenth, 2024; Wickham et al., 2019, 2023). Across models, a random intercept for participant was included to account for individual differences. In cases where a model did not converge, the random intercept was removed.

#### Research Question 1: Across three group of participants (young neurotypical, age-matched neurotypical, individuals with aphasia), are there differences in cortical activity for narrative production vs. counting aloud in bilateral fronto-temporoparietal ROIs as measured via fNIRS?

Estimated oxygenated hemoglobin (HbO) and deoxygenated hemoglobin (HbR) were binned into averaged 3-second intervals (Epoch 1: -2 to 1 second; Epoch 2: 1-4 seconds; Epoch 3: 4-7 seconds; Epoch 4: 7-10 seconds; Epoch 5: 10-13 seconds; Epoch 6: 13-16 seconds; Epoch 7: 16-19 seconds). A separate mixed effects model was fit to predict HbO and HbR for each of the eight ROIs with predictors being the condition x group interaction and all lower terms of that interaction. Epoch was included as a covariate. Average talk time per condition for each participant was included as a covariate to account for differences in response length by individual and condition. Significant interactions were followed up with pairwise comparisons using the emmeans packages with Tukey’s method for multiple comparison correction (Lenth, 2024).When the condition x group interaction was not significant, it was removed from the model and the model was refit. FDR correction for multiple comparisons (Benjamini & Hochberg, 1995) was completed across models within a chromophore (i.e., correction for HbO based upon all p-values for covariates of interest within the HbO models). Given the preliminary nature of the study, both uncorrected and corrected results are reported.

#### Research Question 2 In three groups (young neurotypical, age-matched neurotypical, individuals with aphasia), are there relationships between cortical activity during narrative production and in-task behavioral performance in bilateral fronto-temporoparietal ROIs?

For each group separately, a correlation matrix was generated relating linguistic measures based upon in-task performance to averaged HbO from 5-15 seconds in each of the eight ROIs for the narrative condition. For each ROI with significant correlations to behavioral data, behavioral metrics were input into a backward stepwise regression predicting HbO. Non-significant predictors were removed in a stepwise fashion starting with the predictor with the highest p-value until all predictors were significant at an alpha level of .05.

## 3. Results

### 3.1 Behavioral assessment and behavioral performance

Individuals with aphasia had an average WAB-R AQ score of 82.8 (SD = 15.7) ranging from 54.1 (moderately impaired) to 100 (not aphasic by WAB-R). See Table 4 for standardized test scores. For follow-up yes/no comprehension questions, the young neurotypical group’s average accuracy was 98% (Range: 90–100%); the age-matched group’s average accuracy was 94% (Range: 85–100%); and the aphasia group’s average accuracy was 88% (Range: 65– 100%), indicative of generally good basic comprehension of the story across groups.

### 3.2 fNIRS results

Full statistical output for HbO and HbR and results for HbR analysis are reported in the Supplemental Material. See Table 5 for summary of sample size for each ROI and each group after channel loss due to lesioned tissue in people with aphasia. See Table 6 for brief summary of Research Question 1 HbO results.

**Table 5:** Summary of sample size for each ROI after channel loss.

|  | Left Hemisphere |  |  |  | Right Hemisphere |  |  |  |
|---|---|---|---|---|---|---|---|---|
| Group | Frontal | Temporal | Parietal | Precentral | Frontal | Temporal | Parietal | Precentral |
| Young | 14 | 14 | 14 | 14 | 14 | 14 | 14 | 14 |
| Age-Matched | 15 | 15 | 15 | 15 | 15 | 15 | 15 | 15 |
| PWA | 12 | 10 | 11 | 12 | 17 | 17 | 17 | 17 |

**Table 6:** Summary of Research Question 1 HbO Results. Summary of Research Question 1 significant simple effects of epoch, condition, group, and group x condition interactions. *Survives multiple comparison correction; PWA: People with aphasia; N: narrative condition; C: control condition. NS: not significant. In Research Question 1, the group x condition interaction in the left temporal ROI showed a condition effect (narrative > counting) in the young group not seen in the age-matched group. The group x condition interaction in the right frontal ROI, showed condition effects (narrative > counting) in the young and aphasia groups not seen in the age-matched group. Follow-up pairwise comparisons for a left precentral interaction were not significant after correction.

| Left |  |  |  |  |
|---|---|---|---|---|
| ROI | Frontal | Temporal | Parietal | Precentral |
|  | Condition: N > C*<br>Group: NS<br>Interaction: NS<br>Epoch: 3*,5,6*,7* > 1 | Interaction: Condition x Group<br>Epoch: 5,6*,7* > 1 | Condition: NS<br>Group: NS<br>Interaction: NS<br>Epoch: 6,7* > 1 | Interaction: Condition x Group*<br>Epoch: 5*,6*,7* > 1 |
| Right |  |  |  |  |
| ROI | Frontal | Temporal | Parietal | Precentral |
| Research Question 2 |  |  |  |  |
|  | Interaction: Condition x Group*<br>Epoch: 3*,4,5*,6*,7* > 1 | Condition: N > C*<br>Group: NS<br>Interaction: NS<br>Epoch: 2,3*,4,5*,6*,7* > 1 | Condition: NS<br>Group: NS<br>Interaction: NS<br>Epoch: 2*,3*,6*,7* > 1 | Condition: NS<br>Group: NS<br>Interaction: NS<br>Epoch: 2*,3*,5*,6*,7* > 1 |

#### 3.2.1 Research Question 1: Across the three group of participants (young neurotypical, age-matched neurotypical, individuals with aphasia) are there differences in cortical activity for narrative production vs. counting aloud in bilateral fronto-temporoparietal ROIs as measured via fNIRS?

##### 3.2.1.1 Uncorrected results

Group and Condition:

Primary ROIs: Before multiple comparison correction, in the left frontal ROI there was a simple effect of condition (narrative > counting; *p < .001*). In the left temporal ROI, there was a significant group (young vs. age-matched) by condition interaction (*p = .039*). Follow-up pairwise comparison showed that the young neurotypical group had greater HbO for narrative vs. counting (adjusted *p = .005*) that was not seen in the age-matched group (adjusted *p = .91*). In the right frontal ROI, there were significant group (young vs. age-matched and PWA vs. age-matched) by condition interactions (both *p < .05*). Follow-up pairwise comparison showed greater HbO for narrative vs. counting for the young neurotypical group (adjusted *p < .001*) and the individuals with aphasia (adjusted *p < .001*) that was not seen in the age-matched group (adjusted *p = .646*). In addition, there was greater HbO for narrative in the individuals with aphasia than the age-matched group (adjusted *p = .027*). In the right temporal ROI, there was a simple effect of condition (narrative > counting; *p = .007*).

Exploratory ROIs: There were no significant interactions or effects of group or condition in bilateral parietal ROIs (all unadjusted *p > .05*).

Control ROIs: In the left precentral (control) ROI, there was a significant group (PWA vs. age-matched) x condition interaction (*p = .018*). Although follow-up pairwise comparison did not show any significant differences after multiple comparison correction (all adjusted *p > .05*), the trend was toward individuals with aphasia showing greater cortical activity for counting than narrative.

Epoch: There were simple effects of epoch (with later epochs having greater HbO than Epoch 1) to some extent in all ROIs (all *p < .05*): Ieft frontal (Epoch 3 > 1, 5 > 1, 6 > 1, 7 > 1); left temporal (Epoch 5 > 1, 6 > 1, 7 > 1); right frontal (Epoch 3 > 1, 4 > 1, 5 > 1, 6 > 1, 7 > 1); right temporal (Epoch 2 > 1, 3 > 1, 4 > 1, 5 > 1, 6 > 1, 7 > 1); left parietal (Epoch 6 > 1, 7 > 1); right parietal (Epoch 2 > 1, 3 > 1, 6 > 1, 7 > 1); left precentral (Epoch 5 > 1, 6 > 1, 7 > 1); and right precentral (Epoch 2 > 1, 3 > 1, 5 > 1, 6 > 1, 7 > 1).

##### 3.2.1.2 Corrected results

Group and Condition: After multiple comparison correction, there remained a simple effect of condition (narrative > counting) in the left frontal and right temporal ROIs (all adjusted *p < .05*). There also remained significant condition by group interactions in the right frontal and left precentral ROIs (all adjusted *p < .05*).

Epoch: There remained simple effects of epoch (with later epochs having greater HbO than Epoch 1) to some extent in all ROIs (all adjusted *p < .05*): Ieft frontal (Epoch 3 > 1, 6 > 1, 7 > 1); left temporal (Epoch 6 > 1, 7 > 1); right frontal (Epoch 3 > 1, 5 > 1, 6 > 1, 7 > 1); right temporal (Epoch 3 > 1, 5 > 1, 6 > 1, 7 > 1); left parietal (Epoch 7 > 1); right parietal (Epoch 2 > 1, 3 > 1, 6 > 1, 7 > 1); left precentral (Epoch 5 > 1, 6 > 1, 7 > 1); and right precentral (Epoch 2 > 1, 3 > 1, 5 > 1, 6 > 1, 7 > 1). (See Table 6 and Figure 3 for summary of results).

**Figure 3:**
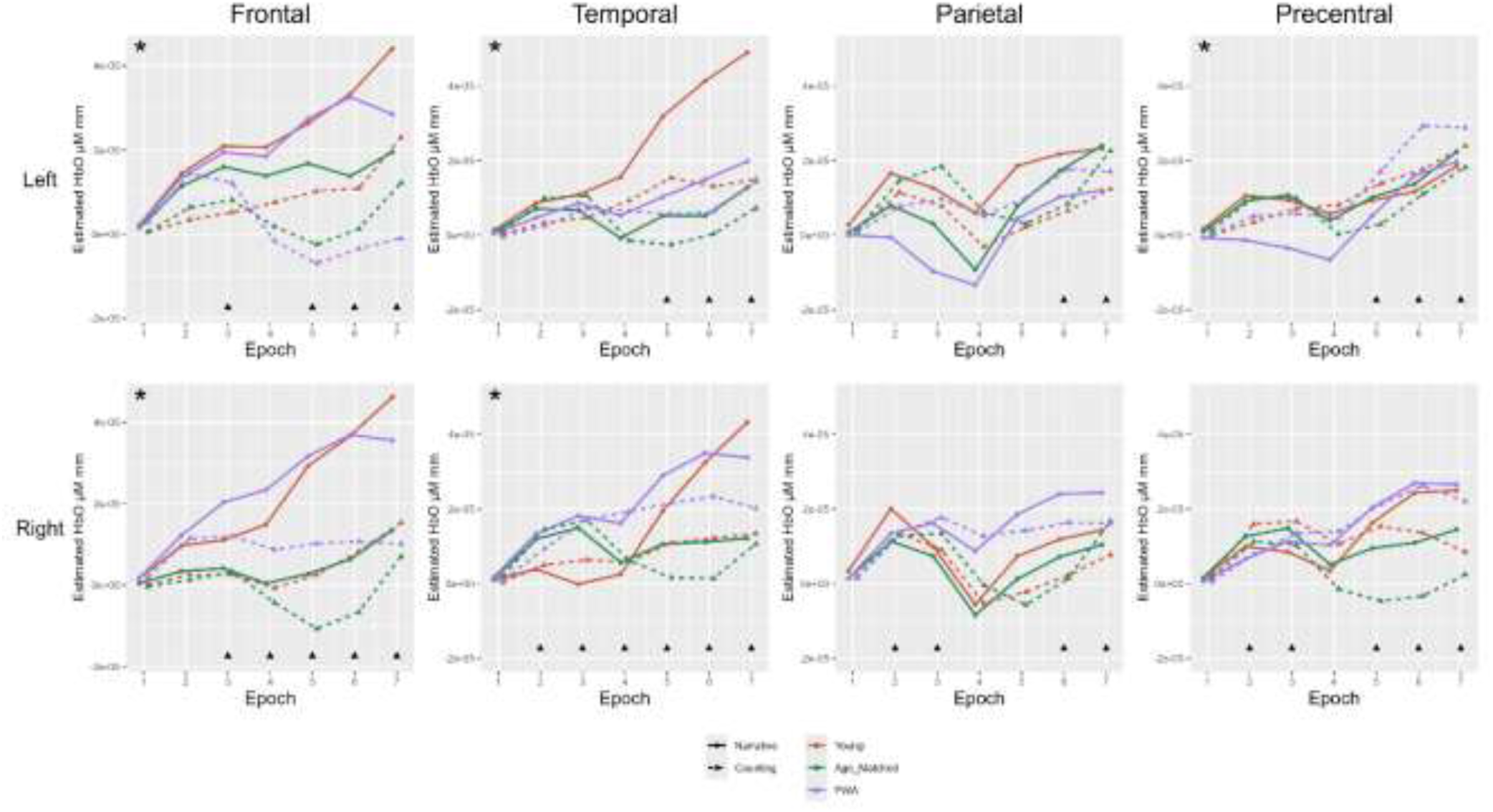
HbO across groups and conditions. Research Question 1 results, with solid lines indicating narrative condition and dashed lines indicating counting condition. The young neurotypical group is represented in red, the age-matched group in green, and the people with aphasia in purple. *indicates significant effect of condition (narrative > counting (left frontal and right temporal) or significant group x condition interaction (where narrative > counting in a subset of groups). For the left temporal ROI, the young neurotypical group showed a condition effect (narrative > counting) not seen in the age-matched group. For the right frontal ROI, the young neurotypical group and people with aphasia showed condition effects (narrative > counting) not seen in the age-matched group. Follow-up pairwise comparisons for a left precentral interaction were not significant. ▴ indicates significant effect of epoch (as compared to Epoch 1).

#### 3.2.2 Research Question 2: In three groups (young neurotypical, age-matched neurotypical, individuals with aphasia), are there relationships between cortical activity during narrative production and in-task behavioral performance in bilateral fronto-temporoparietal ROIs?

Exploratory ROIs: In the young neurotypical group in the left parietal ROI, mean length of utterance in morphemes was a positive predictor of HbO (i.e., longer utterances length was associated with greater HbO; *p = .003*; R^2^ = .53; See Figure 4a).

**Figure 4:**
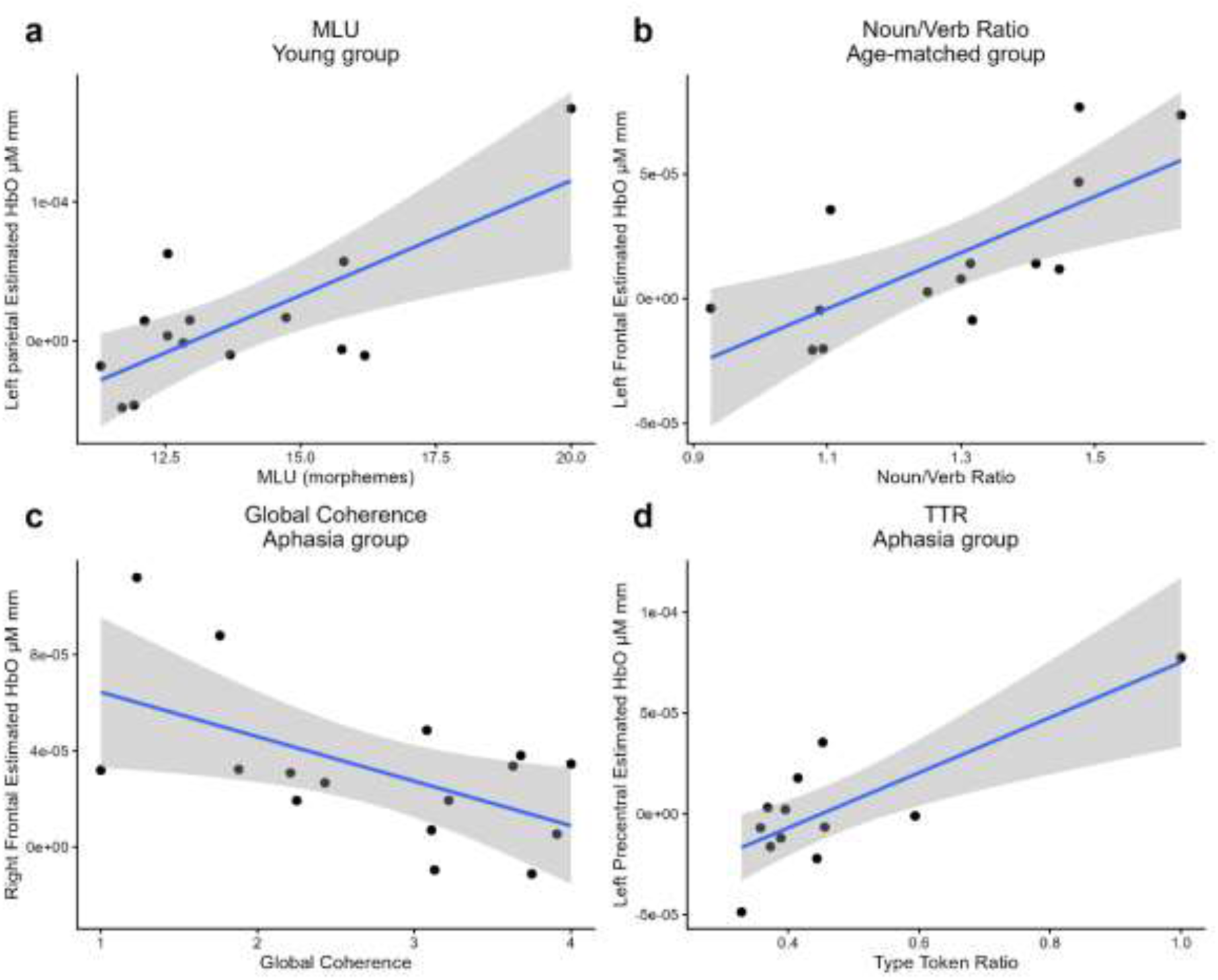
Relationships between discourse measures and HbO.

Primary ROIs: In the age-matched group in the left frontal ROI, noun-verb ratio was a positive predictor of HbO (*p = .004*; R^2^ = .52). In other words, individuals with a higher noun-verb ratio (proxy for less complex sentences) tended to show greater HbO (See Figure 4b).

In the people with aphasia in the left precentral ROI, type-token ratio (a proxy for lexical diversity) was a positive predictor of HbO (*p = .002*; R^2^ = .63, i.e., participants with higher type token ratio tended to show greater HbO; See Figure 4c). In the people with aphasia in the right frontal ROI, global coherence was a negative predictor of HbO (*p = .024*; R^2^ = .32; i.e., participants with lower average global coherence tended to show greater HbO; See Figure 4d).

## 4. Discussion

### 4.1 Overall Summary

Overall, this study showed bilateral frontotemporal cortical activity during narrative production when evaluated relative to counting aloud, with the strongest effects in young individuals. Across ROIs, cortical activity increased over the course of the response periods, reflective of task-based cortical engagement. For the left frontal and right temporal ROIs, narrative production resulted in significantly greater cortical activity than counting, without statistical support for between-group differences. In the left temporal ROI, narrative production resulted in significantly greater cortical activity than counting aloud for the young neurotypical, with neither the age-matched group nor the individuals with aphasia showing a significant effect. In the right frontal ROI, narrative production resulted in significantly greater cortical activity than counting aloud in the young group and the individuals with aphasia, but not the age-matched group. For control ROIs (bilateral precentral gyri), there were not group and condition effects in the right precentral gyrus. However, there was evidence in the left precentral gyrus in the individuals with aphasia of a trend toward greater activity for counting aloud than narrative.

### 4.2 Research Question 1: Across three group of participants (young neurotypical, age-matched neurotypical, individuals with aphasia), are there differences in cortical activity for narrative production (experimental task) vs. counting aloud (control task) in bilateral fronto-temporoparietal ROIs as measured via fNIRS?

For the primary ROIs, these results are broadly consistent with initial hypotheses and prior literature implicating a left-lateralized network of cortical regions needed for the complex task of speech and language processing in young, healthy individuals (Hickok & Poeppel, 2007; Price, 2012). Specifically, the condition effects for HbO in bilateral frontotemporal ROIs are consistent with the hypothesis that participants would show greater responses for the narrative condition across ROIs given the greater linguistic demand of narrative production. These results also confirm the suitability of fNIRS for capturing task-based cortical activity for verbal narrative discourse tasks and provide support for the contrast validity of this novel task.

In the age-matched group, the more limited condition-related effects in the primary ROIs were unexpected. These could be due to generally reduced cortical activity with aging. There is some prior literature to suggest that older adults display under-recruitment of neural resources during cognitive tasks (Maillet & Rajah, 2014); however, there are also a number of studies showing increased cortical activity in this population potentially reflective of neural compensation (Maillet & Rajah, 2014; McDonough et al., 2022). Further study with a larger sample size would help determine if the results in this study are anomalous.

In the primary ROIs in the individuals with aphasia, results were consistent with hypotheses, with greater HbO for narrative vs. counting in bilateral frontal and right temporal ROIs. This could be due to a reliance on right hemisphere homologues of damaged regions. This is consistent with modest prior evidence of recruitment of right-hemisphere homologues of left-hemisphere language regions during language tasks in people with aphasia in fMRI literature (Kiran et al., 2019; Schneck et al., 2026; Wilson & Schneck, 2021). Another possibility is that left temporal effects could not be detected due to small sample size, given that ROIs falling within lesioned tissue were excluded. It may be the case that with a larger sample size, a similar significant pattern would be seen in the left temporal ROI for people with aphasia.

In the control ROIs (bilateral precentral gyri), results were largely consistent with hypotheses that there would not be a significant difference between conditions. However, the finding of a trend toward greater cortical activity for counting vs. narrative in the individuals with aphasia was unexpected. One possibility is that, unlike the neurotypical individuals, the people with aphasia were producing more content in the counting condition leading to greater articulatory effort and precentral activity.

### 4.3 Research Question 2: In three groups (young neurotypical, age-matched neurotypical, individuals with aphasia), are there relationships between cortical activity during narrative production and in-task behavioral performance in bilateral fronto-temporoparietal ROIs?

For Research Question 2, relationships between cortical activity and in-task behavioral measures were explored to evaluate whether and how the patterns of cortical activity seen in Research Question 1 were supporting task performance across groups. The young neurotypical group showed a positive relationship between mean length of utterance and HbO in the left parietal ROI (i.e., greater HbO in the left parietal ROI was related to longer mean length of utterance). This is consistent with the hypothesis that language performance would be positively related to greater cortical activity across groups. This is also broadly consistent with theory and extensive fMRI evidence for left-hemisphere lateralization of language (Hickok & Poeppel, 2007). A lack of relationships for other measures may be due to a ceiling effect in some cases. For example, in the case of global coherence scores, where the scale ranges from 1-4, across the board young neurotypical individuals tended to produced discourse rated at 4.

The age-matched group showed a relationship in the left frontal ROI in which higher noun-verb ratio (proxy for less syntactically complex sentences) were related to greater HbO. The inverse relationship between cortical activity in the left frontal ROI and syntactic complexity in the age-matched group is unexpected. One possibility is that left frontal HbO is indexing linguistic demand as well as cognitive effort, with greater cortical activity in participants with less complex output reflective of greater cognitive effort for task completion given the prefrontal cortex’s involvement in managing cognitive demands (e.g., Shirzadi et al., 2025).

The people with aphasia showed a) a relationship in the left precentral ROI in which higher type-token ratio (proxy for lexical diversity) was related to greater HbO; and b) a relationship in the right frontal ROI in which higher global coherence was related lower HbO. The relationship between left precentral cortical activity and type-token ratio in the people with aphasia is consistent with prior research showing greater left-hemisphere cortical activity positively linked to productive language performance in people with aphasia (Wilson & Schneck, 2021). The inverse relationship between cortical activity and global coherence in the individuals with aphasia in the right frontal ROI is unexpected. Post-hoc tests show that global coherence was correlated both with Western Aphasia Battery Aphasia Quotient (WAB-R AQ) and mean length of utterance in the people with aphasia (i.e., individuals with lower global coherence scores in the discourse task also tend to be those with shorter utterances and more severe aphasia). This may be reflective of right hemisphere recruitment, consistent with some prior evidence of increased recruitment for language processing in general in those with larger lesions and more severe aphasia (Kiran et al., 2019).

### 4.4 Limitations and Future Directions

Some limitations of this study should be mentioned. First, there were confounds in the experimental contrast that likely influenced results. Variable response times for the narrative vs. the counting condition could have contributed to differences in cortical responses. This was mitigated by incorporating average talk time per condition at the participant level as a regressor into statistical models. In addition, the control condition was inherently less difficult than the experimental condition for neurotypical participants, which makes it difficult to know if cortical activity differences between conditions is reflective of language-specific effort or domain-general cognitive demand in these groups. This is a pervasive issue in neuroimaging research and particularly difficult to solve when examining narrative production.

Second, subject-level digitization of optode locations was not completed due to inconsistency of the 3D digitization equipment available to the investigators. Future work should incorporate digitization for more accuracy in verifying subject-specific ROIs.

Third, an additional consideration is potential between-group differences other than age. All groups were convenience samples and the young neurotypical group in particular was more racially and ethnically diverse than the individuals with aphasia and age-matched controls. The racial and ethnic composition of the young neurotypical individuals was reflective of the university population from which they were recruited. This is of particular note for fNIRS research given that hair and skin pigmentation can differ between these groups and darker hair color in particular can lead to reduce signal quality (Yücel et al., 2025). Care was taken to maximize signal quality during data acquisition. In addition, data channels with poor signal quality were excluded by the data processing stream. However, future work should be mindful of matching groups on extraneous factors that may influence between-group comparisons.

Future work could explore more sophisticated task design with a) attempts to balance domain-general cognitive difficulty across conditions; and b) adjustments to task difficulty based upon severity of verbal expression impairment for people with aphasia. This would help control domain-general cognitive demand in order to better examine cortical underpinnings of narrative language production specifically. In addition, future work could explore cortical activity for discourse production across contexts. For example, different types of monologic discourse production (i.e., procedural, expositional) could be evaluated given the varying linguistic demands of these tasks and potentially different underlying cortical activity needed to support these tasks. Furthermore, evaluation of discourse production in increasingly naturalistic contexts could provide insight into the underlying neural activity supporting communication in real-world situations.

## Supporting information

Manuscript v2.2

## Data Availability

Data and statistical analysis code used in this study are available in Open Science Framework: https://osf.io/q9utc/overview?view_only=9646ccbb70644d15be02c45e35b301c1

## Declaration of Generative AI use

ChatGPT was used for R code optimization. Generative AI was not otherwise used in preparation of this manuscript.

## Data Statement

Raw data and statistical analysis code for this study are available in on Open Science Framework.

## Acknowledgements

The authors thank Natalie Gilmore for training team members in fNIRS methodology; Fatemeh Abdollahi, Michael Scimeca, and Manuel Marte for contributions to task design; Nelson Leonardo Gonzalez Dantas and Kylie Isenburg for task programming; Nishaat Mukadam and Ollie Combs for project management; Grace Mooradian for stimuli recording; Kassidy Bate for stimuli editing; Natalie Peterman, Saicharan Kasetti, Yashvi Grover, Erin Shivers, Madeline Silverstein, Grace Nesbitt, Makenzie Meier, and Nicole Tonyan for language sample transcription; Sreekanth Kura for fNIRS cap printing and welding; Boston University Research Computing Services for consultation on R code optimization; Anne Billot and Isaac Falconer for lesion drawing; Alexander von Lühmann for grant conceptualization; Shalom Henderson for consultation on statistical analysis; and Tyler Perrachione and Sofia Vallila Rohter for feedback as part of the first author’s dissertation committee.

## Author Contributions (CRediT)

Emily J. Braun: Formal analysis, Investigation, Data curation, Writing – original draft, Writing – Review and Editing, Visualization

Erin A. Carpenter: Formal analysis, Investigation

Yuanyuan Gao: Methodology, Software, Investigation, Writing – review and editing Meryem Yücel: Conceptualization, Methodology, Software, Writing – review and editing, Supervision, Funding acquisition

David A. Boas: Conceptualization, Methodology, Software, Resources, Writing – review and editing, Supervision, Funding acquisition

Swathi Kiran: Conceptualization, Methodology, Resources, Writing – original draft, Writing – review and editing, Supervision, Project administration, Funding acquisition

