## Supplementary material for "Cortical activity during narrative discourse production in individuals with post-stroke aphasia and controls measured via functional near-infrared spectroscopy": Manuscript v2.2

**Table 4**: Standardized Test Scores for Individuals with Aphasia

|  | WAB | | CLQT+ | | | | | | BNT | ASRS | SAQOL | CETI |
| --- | --- | --- | --- | --- | --- | --- | --- | --- | --- | --- | --- | --- |
| ID | AQ | Aphasia Subtype | Composite | Attention | Memory | Executive | Language | Visuospatial | % | Raw | Communication | Avg |
| A1 | 89.9 | Anomic | 3.8 | 4 | 3 | 4 | 4 | 4 | 95% | 0 | 2.71 | 56 |
| A2 | 97.8 | Not aphasic* | 4.0 | 4 | 4 | 4 | 4 | 4 | 92% | 6 | 4.71 | 74 |
| A3 | 90 | Anomic | 3.2 | 4 | 2 | 4 | 2 | 4 | 83% | 2 | 2.00 | 56 |
| A4 | 95.3 | Not aphasic* | 3.6 | 4 | 3 | 4 | 3 | 4 | 95% | 2 | 4.71 | - |
| A5 | 72.5 | Conduction | 3.6 | 4 | 4 | 3 | 3 | 4 | 30% | 3 | 3.86 | 62 |
| A6 | 54.1 | Broca's | 2.4 | 3 | 2 | 2 | 1 | 4 | 10% | 25 | 3.14 | 61 |
| A7 | 91.4 | Anomic | 3.8 | 4 | 4 | 4 | 3 | 4 | 87% | 1 | 4.00 | - |
| A8 | 93.2 | Anomic | 4.0 | 4 | 4 | 4 | 4 | 4 | 87% | 2 | 3.71 | 73 |
| A9 | 96.4 | Not aphasic* | 4.0 | 4 | 4 | 4 | 4 | 4 | 97% | 6 | 4.71 | 92 |
| A10 | 55.8 | Broca's | 2.8 | 3 | 2 | 3 | 2 | 4 | 68% | 23 | 2.57 | 57 |
| A11 | 96 | Not aphasic* | 3.4 | 4 | 3 | 3 | 3 | 4 | 83% | 2 | 4.57 | - |
| A12 | 94.9 | Not aphasic* | 3.2 | 4 | 2 | 4 | 2 | 4 | 80% | 5 | 2.86 | 40 |
| A13 | 72.1 | Wernicke's | 3.2 | 4 | 2 | 4 | 2 | 4 | 87% | 2 | 2.71 | 65 |
| A14 | 100 | Not aphasic* | 4.0 | 4 | 4 | 4 | 4 | 4 | 98% | 0 | 4.71 | 98 |
| A15 | 66.3 | Conduction | 3.2 | 4 | 2 | 4 | 2 | 4 | 33% | 12 | 4.00 | 77 |
| A16 | 80.1 | Anomic | 3.0 | 2 | 4 | 3 | 3 | 3 | 85% | 12 | 2.43 | 43 |
| A17 | 61.7 | Conduction | 3.2 | 4 | 2 | 4 | 2 | 4 | 12% | 6 | 2.57 | - |
| Mean | 82.8 |  | 3.4 | 3.8 | 3.0 | 3.6 | 2.8 | 3.9 | 72% | 6.4 | 3.5 | 65.6 |
| SD | 15.7 |  | 0.5 | 0.6 | 0.9 | 0.6 | 1.0 | 0.2 | 30% | 7.5 | 1.0 | 17.0 |

WAB: Western Aphasia Battery – Revised; AQ: Aphasia Quotient, possible scores range from 0 (severe) to 100 (no impairment; per WAB-R, Aphasia Quotient >93.8 cut-off for aphasia. However, all participants classified as non-aphasic in this study reported functional impairment); CLQT+: Cognitive Linguistic Quick Test, possible Composite and Domain Scores range from 1 (severe) to 4 (within normal limits); BNT: Boston Naming Test, 2^nd^ edition, raw scores /60 converted to a percent accuracy; ASRS – Apraxia of Speech Rating Scale v3.5, possible scores range from 0 (no abnormal speech features) to 52; SAQOL-39 – Stroke and Aphasia Quality of Life Scale – Communication domain average score – self report measure of communicative functioning with possible scores ranging from 1 (“couldn’t do it at all”) to 5 (“no trouble at all”) for several communication situations; CETI – Communicative Effectiveness Inventory average score – a caregiver report measure of functional communication with possible scores for each item ranging from 0 (“not at all able”) to 100 (“as able as before stroke”).
